# Ordinal Multi-Instance Learning for Automated Multi-Task Grading of Lumbar Degenerative Conditions on Sagittal MRI, with External Validation

**DOI:** 10.64898/2026.10.20.26366005

**Authors:** Shijiang Geng, Qizhuo Miao, Qingqing Qin, Haoran Guo, Xin Wang

## Abstract

**Purpose:** Lumbar spine MRI interpretation requires 25 structured severity judgments per examination — five degenerative conditions at five disc levels, each on a three-level ordinal scale — a time-consuming task with substantial inter-reader variability. We developed and externally validated a model that automates this complete scheme.

**Materials and Methods:** A full-sequence multi-instance learning (MIL) model — ResNet34 backbone, bidirectional GRU, attention pooling over 24 uniformly sampled sagittal T2-weighted slices — with 25 cumulative-logit ordinal heads was trained on 1,204 examinations from the multi-institutional RSNA 2024 LumbarDISC dataset and internally validated on 213 held-out examinations. External validation used 60 retrospectively collected patients from an independent Chinese university hospital (four scanner vendors, radiology reports as the reference standard, no fine-tuning). Agreement was assessed with exact accuracy, sensitivity, specificity, McNemar tests, and patient-clustered bootstrap confidence intervals.

**Results:** The ordinal model achieved 51.4% balanced accuracy internally (exact accuracy 65.1%), versus 16.3% for a single-slice baseline; severe-grade AUCs averaged 0.908 across conditions. Externally, it outperformed its softmax-weighted counterpart on exact accuracy (0.649 vs. 0.586; +6.3 percentage points, 95% CI +2.7 to +10.0) and sensitivity (0.868 vs. 0.833), with markedly fewer severe-grade false alarms (69 vs. 172 predicted severe cells; McNemar p<0.0001).

**Conclusion:** A full-sequence MIL model with ordinal outputs automates the complete 25-task grading scheme on sagittal MRI and, in the first hospital external validation of such a model, reduced severe-grade over-prediction under domain shift without loss of sensitivity — supporting its role as a screening and triage aid.

**Summary:** An ordinal full-sequence multi-instance learning model graded all 25 lumbar MRI degeneration tasks and markedly reduced severe-grade false alarms in 60-patient hospital validation without loss of sensitivity.

**Key Points:**

- Full-sequence attention was indispensable: balanced accuracy rose from 16.3% (single-slice baseline) to 51.4% over 24 sagittal slices.
- In external validation, ordinal outputs reduced severe-grade predictions from 172 to 69 of 900 cells (McNemar p<0.0001) with comparable ranking ability (AUC 0.835 vs. 0.829).
- The Moderate grade remained the hardest class across all conditions (AUC 0.70–0.78), tracking known inter-reader variability at the mild–moderate boundary.

## 1. Introduction

Lumbar degenerative disease — spinal canal stenosis, neural foraminal narrowing, subarticular stenosis — is among the most common causes of low back pain and radiculopathy [1,2]. MRI is the reference standard for diagnosis, and examination volumes continue to rise [3], even though degenerative findings are common in asymptomatic individuals [4]. A complete interpretation requires grading five conditions at five disc levels on a three-level ordinal severity scale — 25 judgments per examination — a time-consuming task with substantial inter-reader variability, particularly at the mild–moderate boundary [5]. Automated grading could reduce workload and standardize reporting, provided it generalizes across institutions and vendors.

Deep learning for lumbar MRI began with automated radiological-feature extraction in the Genodisc cohort [6]. Most published models address a single condition — overwhelmingly spinal canal stenosis [7] — with binary or few-class outputs, few input slices, and image-level rather than patient-level evaluation [8]; the strongest current benchmark on the LumbarDISC dataset reports 79.4% balanced accuracy for canal stenosis alone, dropping to 55.4% under direct transfer of pre-trained models [9]. Attention-based multi-instance learning (MIL) weights the informative slices of a variable-length MRI series [10,11] but has rarely been applied to full multi-task spine grading.

The RSNA 2024 LumbarDISC dataset [12] — 1,975 annotated examinations from eight institutions in six countries, spanning five conditions × five levels × three grades — created the first opportunity to benchmark models for the complete scheme. Competition solutions addressing all 25 tasks report balanced accuracies near 50–60%, with the moderate grade the most difficult class [13].

Generalization to clinical data remains a well-documented challenge [14] and is essentially untested for the complete scheme. Two studies externally validated RSNA-derived canal-stenosis models — 70 Thai patients with a custom four-level label scheme [15] and a Bangladeshi cohort [16] — but both cover a single condition with non-standard labels and no patient-level statistics. No model covering all five conditions and five disc levels has been externally validated.

We developed a full-sequence, attention-based MIL model with an ordinal regression formulation for simultaneous grading of all 25 tasks from sagittal T2-weighted MRI, validated internally on 1,417 LumbarDISC examinations and externally on a retrospective, multi-vendor cohort of 60 patients from an independent Chinese university hospital. Contributions: (i) the first hospital external validation of the complete five-condition × five-level scheme, with patient-level statistics (McNemar tests, patient-clustered bootstrap confidence intervals); (ii) evidence that ordinal outputs reduce severe-grade false positives under domain shift (69 vs. 172 predicted severe cells); (iii) a systematic ablation quantifying full-sequence MIL, sagittal input, and inverse-frequency weighting; and (iv) attention analysis showing the model relies on slices away from the sequence midpoint — information single-slice baselines discard.

## 2. Materials and Methods

### 2.1 Dataset

The RSNA 2024 LumbarDISC dataset [12], the largest public collection of annotated lumbar spine MRI (2,697 patients; competition subset 1,975; Table 1), yielded 1,417 patients with both sagittal and axial T2 sequences and complete 25-target annotation, randomly split into training (n=1,204, 85%) and validation (n=213, 15%) sets (fixed seed 42; scikit-learn [17]).

#### Annotation schema

Expert radiologists annotated five conditions — spinal canal stenosis, left/right neural foraminal narrowing, and left/right subarticular stenosis — at five disc levels (L1/L2–L5/S1), yielding 25 targets per study, each graded Normal/Mild, Moderate, or Severe.

#### Label distribution

Of 35,425 annotations, 75.1% were Normal/Mild, 17.6% Moderate, 7.2% Severe; abnormality ranged from 14.2% (canal stenosis) to 31.1% (subarticular stenosis), with L4/L5 most affected (45.6%) (Table 1).

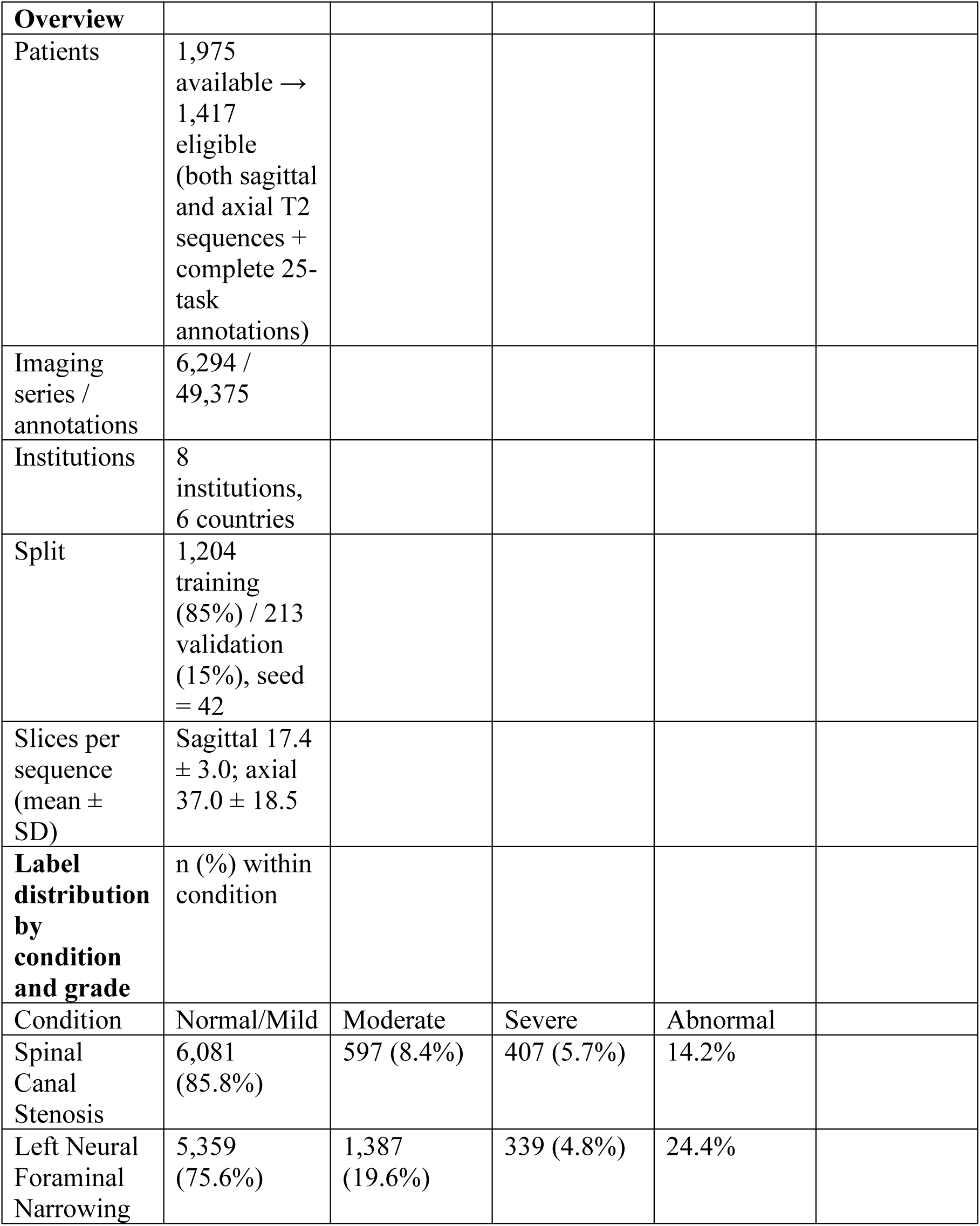
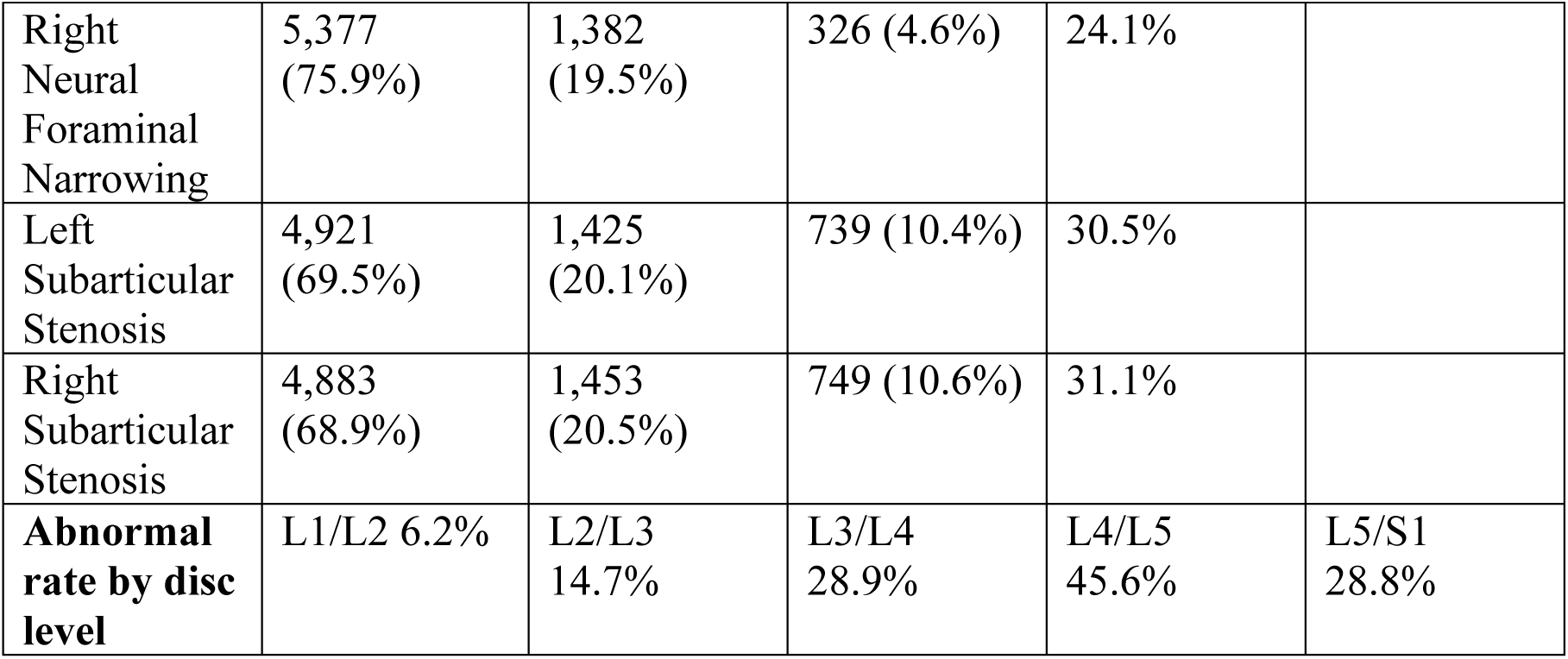

| **Overview** |  |  |  |  |
| --- | --- | --- | --- | --- |
| Patients | 1,975 available → 1,417 eligible (both sagittal and axial T2 sequences + complete 25-task annotations) |  |  |  |
| Imaging series / annotations | 6,294 / 49,375 |  |  |  |
| Institutions | 8 institutions, 6 countries |  |  |  |
| Split | 1,204 training (85%) / 213 validation (15%), seed = 42 |  |  |  |
| Slices per sequence (mean ± SD) | Sagittal 17.4 ± 3.0; axial 37.0 ± 18.5 |  |  |  |
| **Label distribution by condition and grade** | n (%) within condition |  |  |  |
| Condition | Normal/Mild | Moderate | Severe | Abnormal |
| Spinal Canal Stenosis | 6,081 (85.8%) | 597 (8.4%) | 407 (5.7%) | 14.2% |
| Left Neural Foraminal Narrowing | 5,359 (75.6%) | 1,387 (19.6%) | 339 (4.8%) | 24.4% |

|  |  |  |  |  |  |
| --- | --- | --- | --- | --- | --- |
| Right Neural Foraminal Narrowing | 5,377 (75.9%) | 1,382 (19.5%) | 326 (4.6%) | 24.1% |  |
| Left Subarticular Stenosis | 4,921 (69.5%) | 1,425 (20.1%) | 739 (10.4%) | 30.5% |  |
| Right Subarticular Stenosis | 4,883 (68.9%) | 1,453 (20.5%) | 749 (10.6%) | 31.1% |  |
| **Abnormal rate by disc level** | L1/L2 6.2% | L2/L3 14.7% | L3/L4 28.9% | L4/L5 45.6% | L5/S1 28.8% |

### 2.2 Data Preprocessing

DICOM images were parsed with pydicom; the imaging plane was determined from ImageOrientationPatient (IOP) cross-products, cross-checked against the SeriesDescription tag. Each slice was min–max scaled to 8-bit intensity (0–255), resized to 224×224 pixels, converted from grayscale to 3-channel, and normalized with ImageNet statistics. Sagittal T2 sequences were uniformly sampled to 24 slices (axial, 32), with zero-padding or linear-interpolation downsampling as needed. Random horizontal flipping (p=0.5) was applied during training.

### 2.3 Model Architecture

The proposed framework consists of four components (Figure 1):

**Figure 1.**
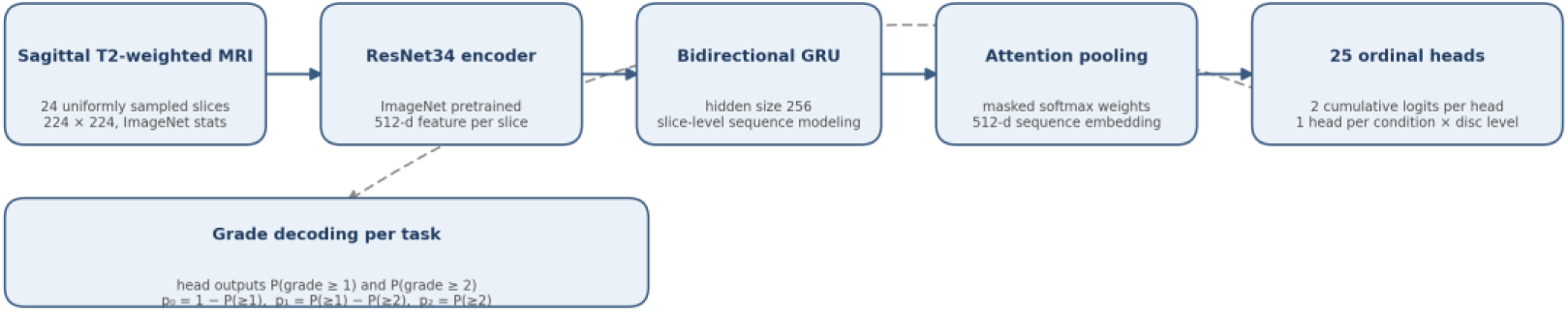
Architecture of the proposed ordinal multi-instance learning framework. Sagittal T2-weighted MRI sequences are uniformly sampled to 24 slices and encoded by a shared ImageNet-pretrained ResNet34 backbone; slice features are aggregated by a bidirectional gated recurrent unit (GRU) followed by masked attention pooling, and the resulting sequence embedding feeds 25 independent cumulative-logit heads (one per condition × disc level), which decode the three ordered severity grades.

#### Shared CNN Encoder

A ResNet34 backbone [18] pretrained on ImageNet [19], with classification and pooling layers removed, output a 512-dimensional feature vector per slice.

#### Bi-directional GRU Sequence Modeling

The slice-feature sequence was processed by a bidirectional gated recurrent unit (GRU) [20] (hidden size 256; 512 after concatenation).

#### Attention-based Pooling

A two-layer perceptron (512→128→1, tanh) generated a scalar attention weight per slice; softmax-normalized weights produced the final sequence representation as the weighted sum of slice features:

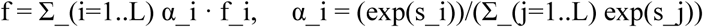

where s_i is the raw attention score and α_i the normalized weight; padded (zero) slices were masked with s_i = -∞.

#### Cross-Plane Attention Fusion (ablation variant)

A dual-plane variant fused sagittal and axial features via multi-head self-attention [21] but did not outperform the sagittal-only model (Section 3.2).

#### Multi-Task Classification Heads

Following the classic multi-task paradigm [22], each of the 25 tasks had an independent three-layer classifier (512→256→ReLU→Dropout(0.3)→128→ReLU→Dropout(0.2)→3) with softmax activation.

### 2.4 Training Protocol

The model was trained for up to 60 epochs with early stopping (patience 15, minimum delta 0.001), AdamW [23] (learning rate 1e-4, weight decay 1e-4) with a cosine warm-restart scheduler, mixed precision (AMP), batch size 4, and gradient clipping (max_norm 1.0).

#### Loss function

Inverse frequency-weighted cross-entropy with label smoothing [24] (ε=0.05); class weights w_c = N_total / (3 × N_c) yielded 0.39 for Normal/Mild, 1.71 for Moderate, and 2.94 for Severe — a standard remedy for class imbalance [25].

#### Ordinal regression (primary model)

The primary model used cumulative-logit ordinal regression [26] implemented as two binary heads per task following the CORAL framework [27], each head estimating a cumulative probability (P(severity ≥ Moderate), P(severity ≥ Severe)); the loss averaged two binary cross-entropy terms. Class probabilities were recovered from the cumulative logits: P(Normal/Mild) = 1 - P(≥Moderate); P(Moderate) = P(≥Moderate) - P(≥Severe); P(Severe) = P(≥Severe).

All experiments ran on a single NVIDIA RTX 4070 Laptop GPU (8 GB VRAM) with PyTorch 2.11 [28] and CUDA 12.8.

### 2.5 Evaluation Metrics

The primary metric was **balanced accuracy** for detecting clinically relevant pathology, defined as:

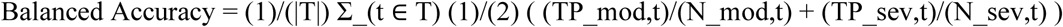

where T is the 25-task set; the metric targets Moderate and Severe detection, excluding the dominant Normal/Mild samples.

Secondary metrics included per-grade one-vs-rest ROC AUC (per-condition and micro-averaged), macro-averaged F1, and confusion matrices.

### 2.6 External Validation

#### Cohort

This retrospective study was approved by the Ethics Committee of the First Hospital of Lanzhou University (approval No. LDYYLL2026-891; 13 October 2026), with written informed consent waived. Sixty patients undergoing lumbar spine MRI for low back or leg pain (February–August 2026) at an independent Chinese university hospital were retrospectively included (mean age 59.3 ± 12.8 years; 32 female; four scanner vendors; Table 4). Only pre-operative examinations were included. Ground truth came from structured radiology reports for the three conditions routinely reported in Chinese practice (spinal canal stenosis and left/right neural foraminal narrowing) at the same five disc levels (15 tasks; subarticular stenosis is not routinely reported). Levels were coded on the training scale (0 = not mentioned/normal or mild; 1 = moderate; 2 = severe); grade 0 thus spans normal findings and unmentioned levels, a convention addressed as a limitation (Section 4.4).

#### Protocol

Both trained models were applied to hospital DICOM studies through the identical preprocessing pipeline (Section 2.2) without fine-tuning; no hospital data were used at any stage of model development; all analyses used anonymized data. One study with an unusually short sagittal acquisition (five slices) was retained via a slice-count fallback and flagged. The first 30 cases were used for model selection and the remaining 30 as a held-out confirmation set; pooled results for all 60 cases are reported.

#### Statistical analysis

Agreement was assessed with exact (three-class) accuracy and, for binary abnormality detection (grade ≥ 1), sensitivity, specificity, and positive predictive value. McNemar’s test [29] with continuity correction compared the two models per condition and overall; 95% confidence intervals for accuracy differences were obtained by patient-clustered bootstrap [30] (2,000 resamples). Discrimination was additionally quantified by cell-level (900 cells) and patient–condition-level AUC with the same bootstrap intervals.

## 3. Results

### 3.1 Baseline Comparison

The single-slice baseline (middle slice only) achieved 16.3% balanced accuracy, marginally above chance; the full-sequence MIL model (sagittal-only, 16 slices) achieved 32.7% — a 101% relative improvement (Table 2).

### 3.2 Ablation Study

#### Input modality

Sagittal-only input (32.7%) outperformed axial-only (26.3%) by 6.4 points, indicating sagittal T2 carries most diagnostic information. The dual-plane variant (29.4%) underperformed by 3.3 points, and mean pooling in place of attention pooling yielded a comparable 30.0%.

#### Loss function

The largest gain came from inverse frequency weighting with reduced label smoothing (0.10 to 0.05); combined with increasing sagittal slices from 16 to 24, balanced accuracy rose from 32.7% to 50.6% (+17.9 points; Table 2, Figure 2A).

**Figure 2.**
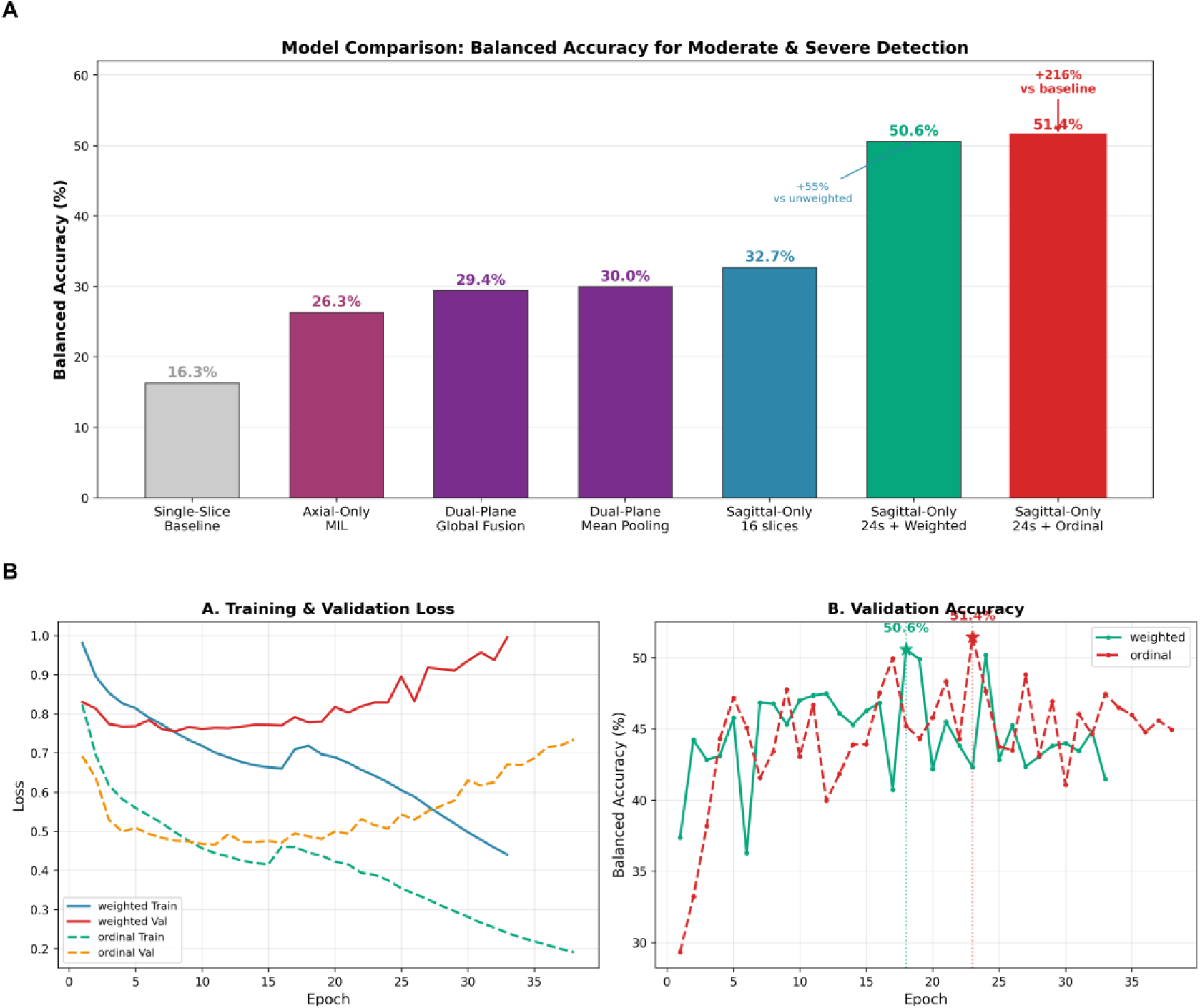
Model development. (A) Ablation comparison: balanced accuracy for detecting moderate and severe pathology across model variants, from the single-slice baseline (16.3%) to the final 24-slice ordinal model (51.4%). (B) Training dynamics of the weighted and ordinal models: training and validation loss, and validation balanced accuracy, with the best epoch marked (ordinal model: epoch 23, 51.4%).

#### Ordinal regression

The ordinal formulation improved balanced accuracy from 50.6% to 51.4% (exact accuracy 65.1%; epoch 23; Figure 2B) and reshaped decision behavior: severe-grade predictions on the validation set fell from 933 to 380 cells, while Moderate-grade AUC rose from 0.721 to 0.756 (+3.5 points) and Severe-grade AUC from 0.914 to 0.917 (Table 3) — a suppression of extreme predictions that proved decisive under domain shift (Section 3.7).

| **Model Variant** | **Balanced Accuracy** |
| --- | --- |
| Single-Slice Baseline | 16.3% |
| Axial-Only MIL | 26.3% |
| Dual-Plane + Cross-Attention | 29.4% |
| Dual-Plane + Mean Pooling | 30.0% |
| Sagittal-Only MIL (16 slices) | 32.7% |
| **Sagittal-Only MIL (24 slices, Weighted Loss)** | **50.6%** |
| **Sagittal-Only MIL (24 slices, Ordinal Regression)** | **51.4%** |

### 3.3 Per-Condition Analysis

AUC varied substantially across conditions (Table 3, Figures 4B and 5A): canal stenosis achieved the highest AUCs (0.932/0.783/0.944), foraminal narrowing the lowest Severe-grade AUCs (0.891 left, 0.876 right). Moderate was universally the most challenging grade, its AUC 11–22 percentage points below the corresponding Severe-grade AUC — consistent with known inter-rater variability at the mild–moderate boundary [5,31].

**Figure 4.**
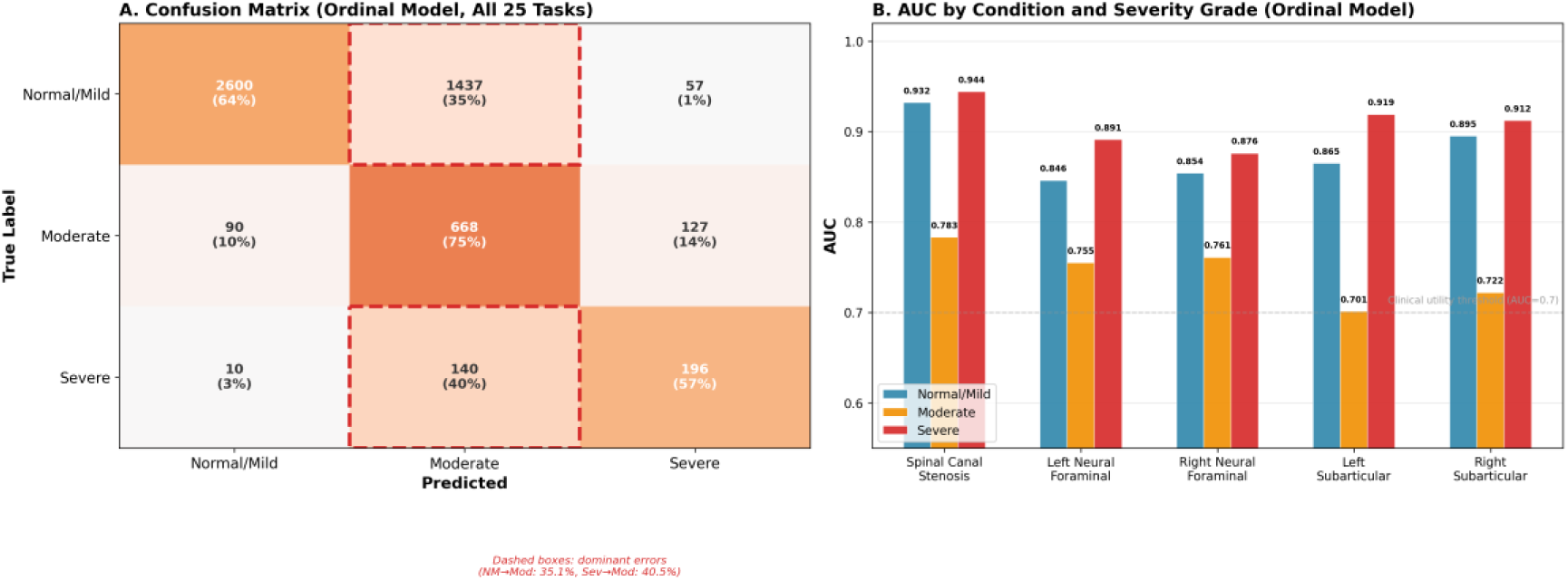
Internal validation performance of the ordinal model. (A) Confusion matrix aggregated over all 25 tasks (5,325 predictions from 213 validation patients); dashed boxes highlight the dominant error directions (Normal/Mild→Moderate, 35.1%; Severe→Moderate, 40.5%). (B) Area under the receiver operating characteristic curve (AUC) by condition and severity grade; the dashed line marks the 0.7 clinical utility threshold.

**Figure 5.**
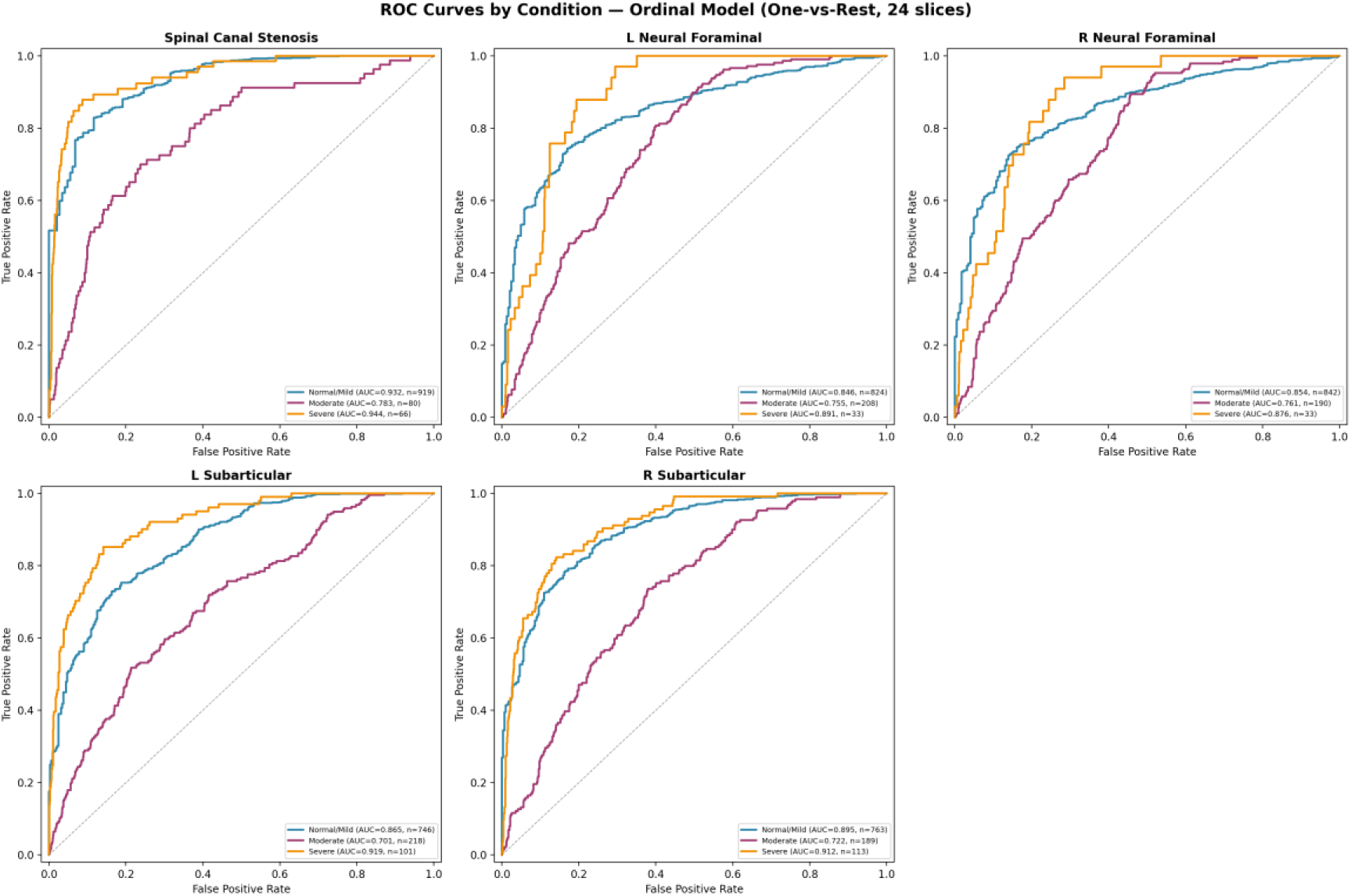
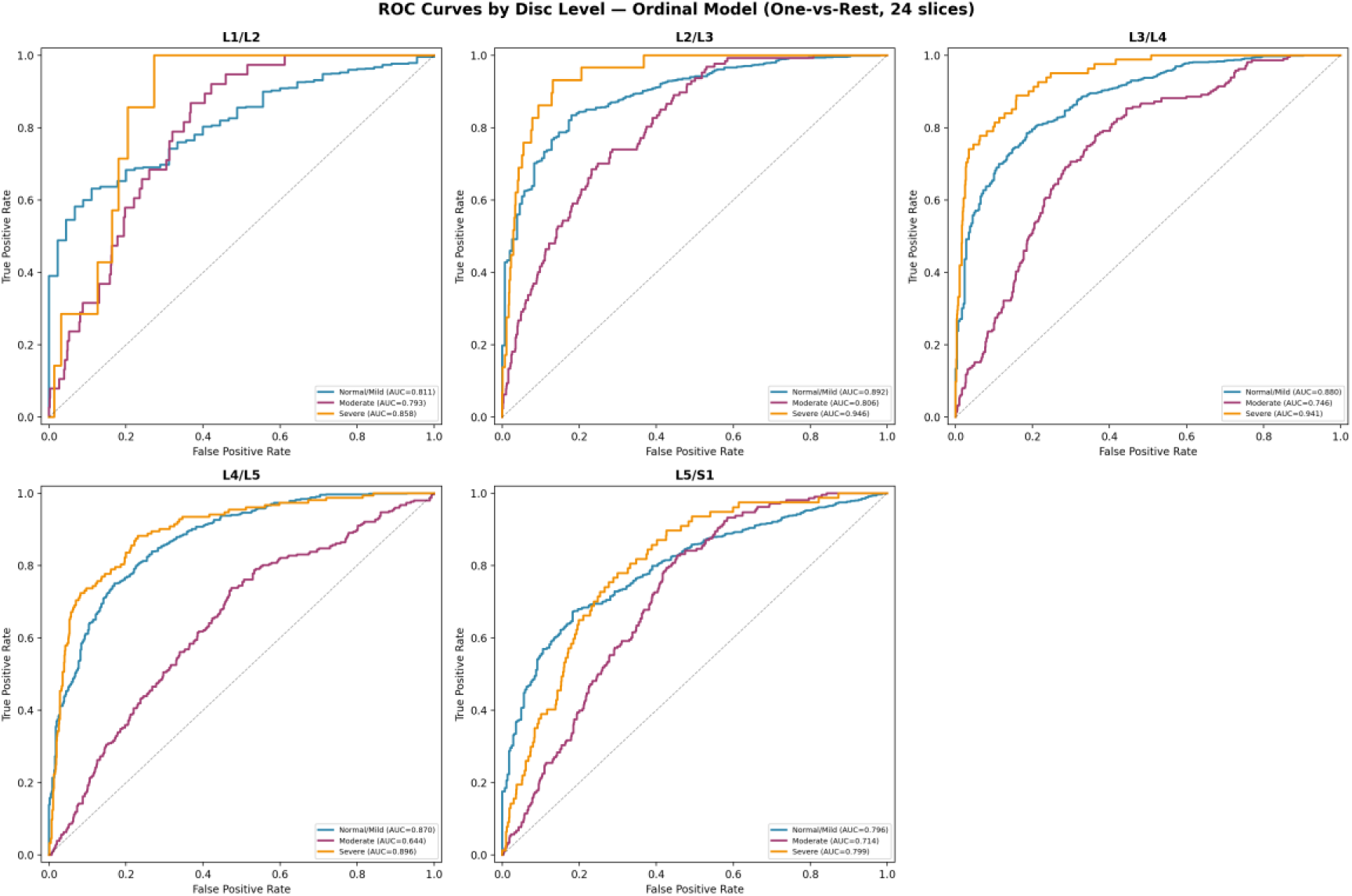
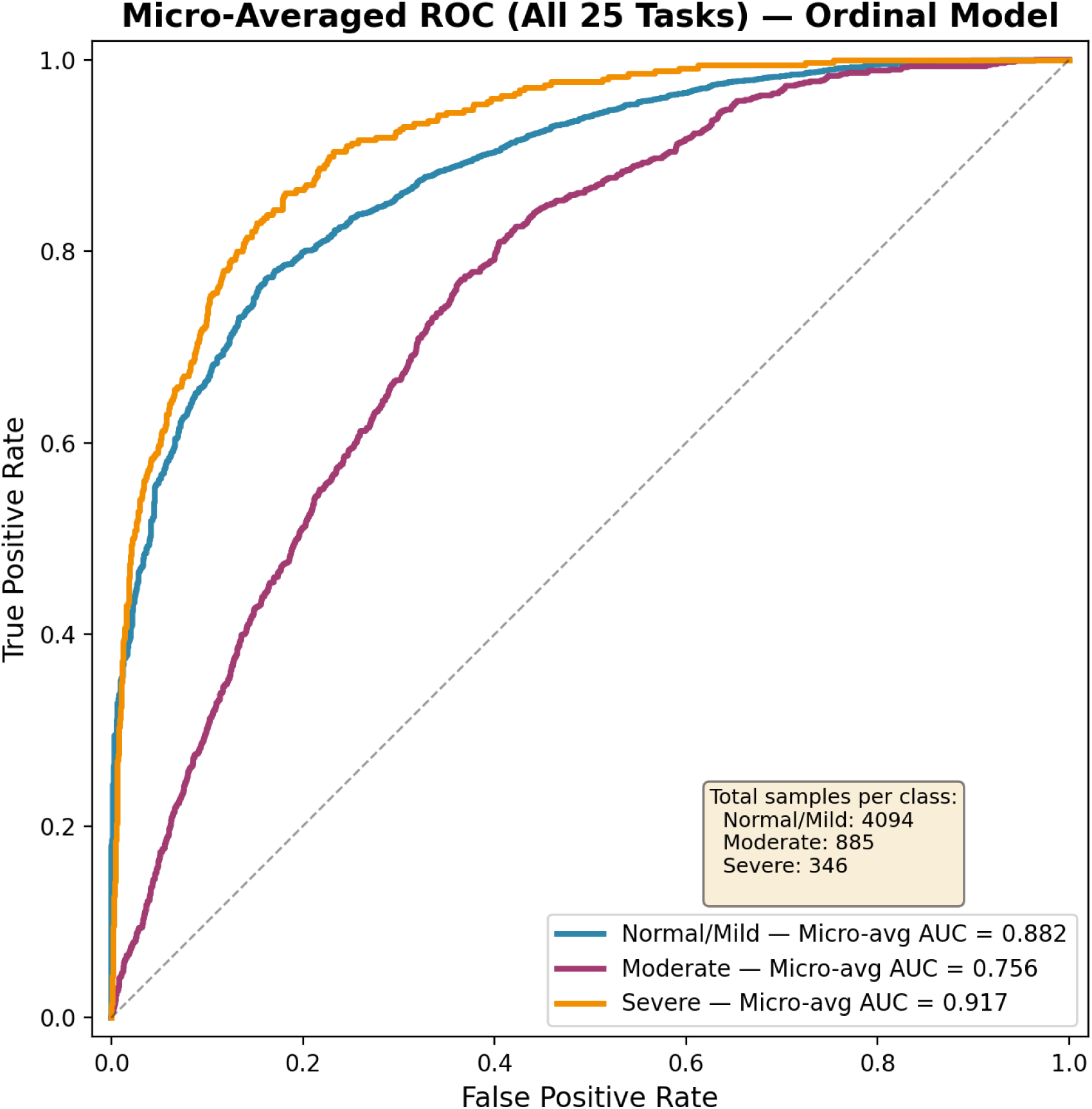
Receiver operating characteristic curves for the ordinal model. (A) By condition; (B) by disc level; (C) micro-averaged across all 25 tasks.

| **Condition** | **Normal/Mild** | **Moderate** | **Severe** |
| --- | --- | --- | --- |
| Spinal Canal Stenosis | 0.932 | 0.783 | 0.944 |
| Left Neural Foraminal Narrowing | 0.846 | 0.755 | 0.891 |
| Right Neural Foraminal Narrowing | 0.854 | 0.761 | 0.876 |
| Left Subarticular Stenosis | 0.865 | 0.701 | 0.919 |
| Right Subarticular Stenosis | 0.895 | 0.722 | 0.912 |
| **Average** | **0.879** | **0.744** | **0.908** |

### 3.4 Per-Level Analysis

Performance tracked disease prevalence across disc levels (Supplementary Table S2, Figure 5B): the most affected levels, L4/L5 (42.5% abnormal) and L3/L4 (27.4%), achieved the highest balanced accuracies (72.6% and 75.6%), while the least affected, L1/L2 (4.2%), reached 26.3%. Prevalence and balanced accuracy correlated strongly (Spearman’s ρ = 0.8, n=5), suggesting sample scarcity at less-affected levels, not model design, is the primary bottleneck.

### 3.5 Confusion Matrix Analysis

Across all 25 tasks and 213 validation patients (5,325 predictions), the ordinal model correctly classified 63.5% of Normal/Mild, 75.5% of Moderate, and 56.6% of Severe labels (Figure 4A). The dominant error was over-prediction of Normal/Mild as Moderate (35.1%); true Severe cases were most often under-called as Moderate (40.5%), and only 2.9% were misclassified as Normal/Mild.

### 3.6 Attention Visualization

Per-slice attention distributions from eight representative validation cases (Figure 6) showed attention concentrated on a few slices (top-3: 47–74% of total weight); in all eight cases the highest-attention slice lay away from the sequence midpoint, with a median displacement of 7 slices (5–8), a positional shift of 30–50% (median 41%) of the sequence. Grad-CAM [32] heatmaps for the same cases are provided in Supplementary Figure S1.

**Figure 6.**
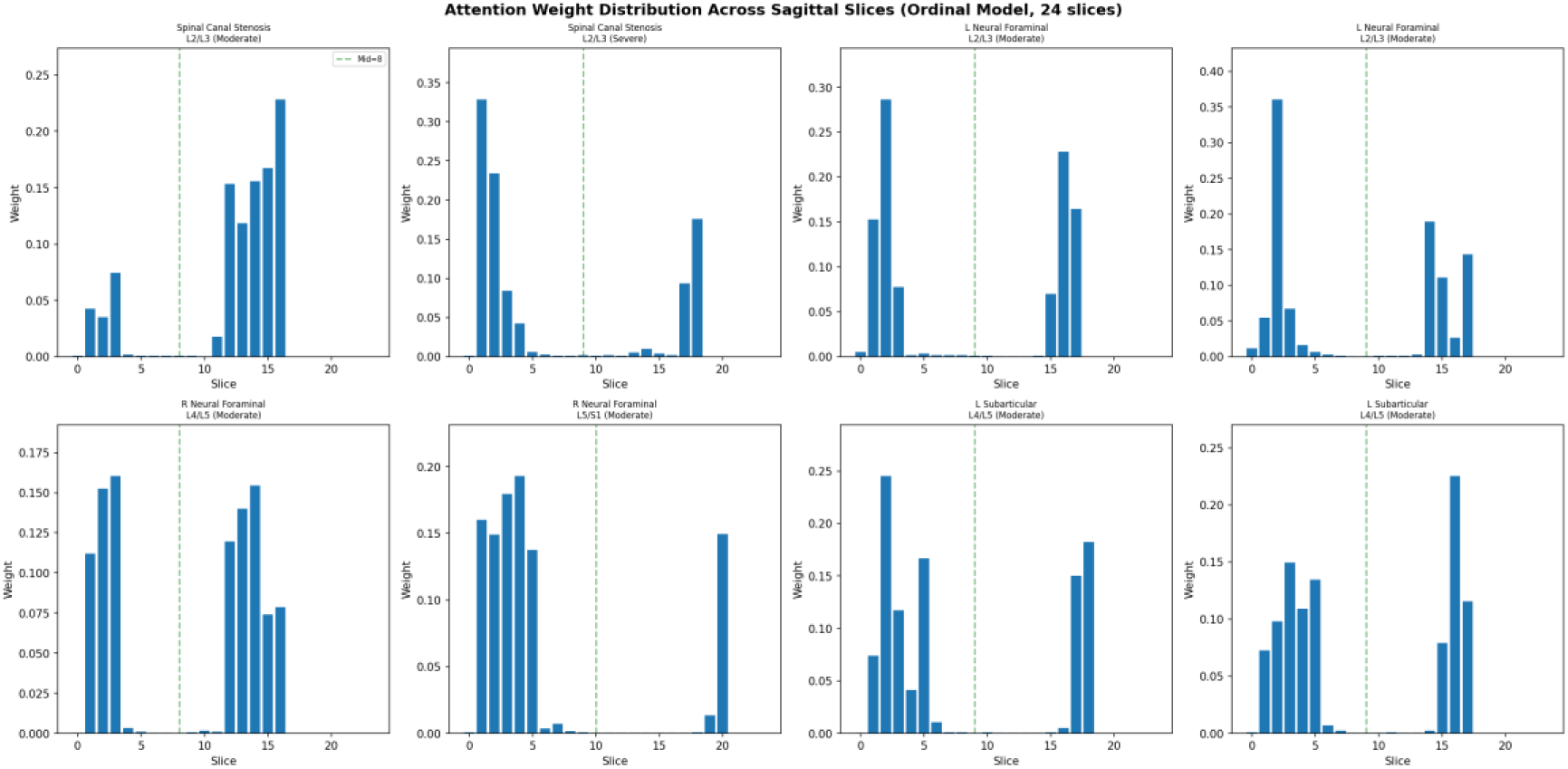
Per-slice attention weight distributions for eight representative validation cases. Attention concentrates on a small subset of slices located away from the sequence midpoint.

### 3.7 External Validation

#### Cohort characteristics

Radiology reports mentioned stenosis in 35 of 60 patients (58.3%). Among 900 patient–level cells, only 7 (0.8%) were graded severe, mirroring the long-tailed source distribution (Table 4).

| Characteristic | Overall (n=60) | Batch 1 (n=30) | Batch 2 (n=30) |
| --- | --- | --- | --- |
| Age (years), mean ± SD | 59.3 ± 12.8 | 59.4 ± 11.5 | 59.2 ± 14.0 |
| Age range | 32–86 | 33–75 | 32–86 |
| Sex, female / male | 32 / 28 | 15 / 15 | 17 / 13 |
| Scanner vendor | Siemens 30, Philips 14, GE 12, United Imaging 4 | Siemens 15, Philips 8, GE 6, United Imaging 1 | Siemens 15, Philips 6, GE 6, United Imaging 3 |
| Report date range | 2026-02-02 to 2026-08-04 | 2026-02-02 to 2026-07-30 | 2026-04-28 to 2026-08-04 |
| Spinal canal stenosis ($\geq 1$ level) | 24 (40.0%) | 14 (46.7%) | 10 (33.3%) |
| Left foraminal narrowing ($\geq 1$ level) | 26 (43.3%) | 12 (40.0%) | 14 (46.7%) |
| Right foraminal narrowing ($\geq 1$ level) | 26 (43.3%) | 13 (43.3%) | 13 (43.3%) |

Note: Batch 1 comprises patients 1–30 (model-selection set) and Batch 2 patients 31–60 (independent test set). Conditions not mentioned in the radiology report were treated as normal (grade 0).

#### Model comparison

The ordinal model achieved higher exact accuracy than the weighted model (0.649 vs. 0.586; +6.3 percentage points, 95% CI +2.7 to +10.0). For binary abnormality detection it yielded accuracy 0.696, sensitivity 0.868, specificity 0.654, and positive predictive value 0.376 (weighted: 0.674, 0.833, 0.636, 0.355). The overall binary McNemar comparison was marginal (p=0.096), driven by canal stenosis, where the ordinal model was significantly better (p=0.0002; 7 vs. 31 discordant cells). The most striking difference concerned severe-grade predictions: 172 cells for the weighted model versus 69 for the ordinal (McNemar p<0.0001). Balanced accuracy favored the weighted model (0.468 vs. 0.395), a reversal entirely attributable to recall on the seven severe ground-truth cells (4/7 vs. 1/7) that should be interpreted with caution given the small severe-grade sample.

#### Discrimination

Cell-level AUC was 0.835 versus 0.829 (Figure 7; CI −0.026 to +0.041), patient–condition-level AUC 0.756 versus 0.799 (CI −0.107 to +0.013), and severe-grade AUC 0.850 versus 0.820. Ranking ability is therefore comparable; the ordinal model’s advantage lies in decision behavior — fewer severe-grade false alarms.

**Figure 7.**
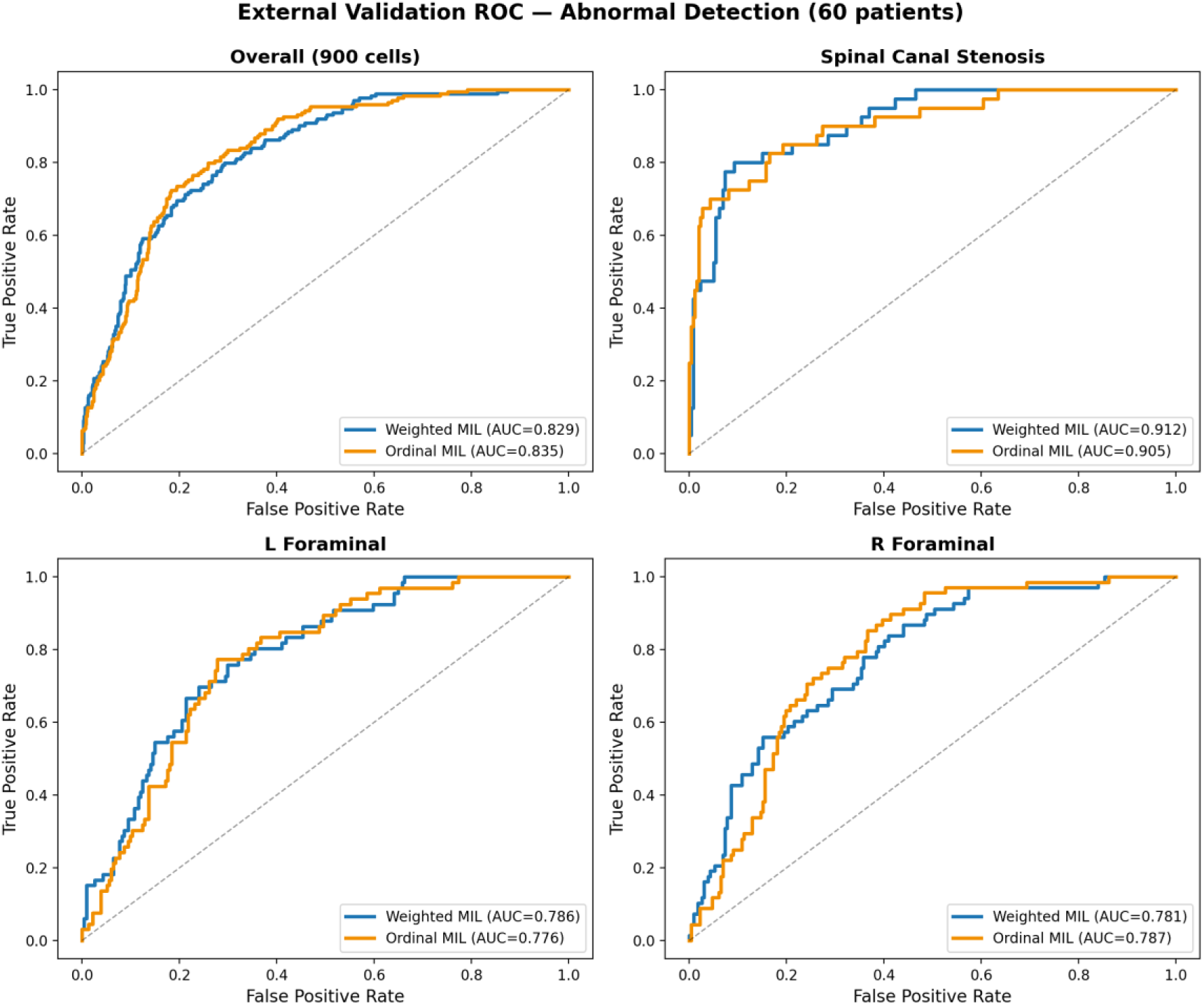
External validation receiver operating characteristic curves for abnormal detection (grade ≥ 1): overall (900 patient-level cells) and by condition, comparing the ordinal and weighted models (60 patients, four scanner vendors).

#### Error analysis

False-negative cells (report-positive, predicted grade 0) numbered 23 for the ordinal model versus 29 for the weighted model (across 14 vs. 16 patients). Among the 25 patients with no mentioned stenosis, the weighted model produced abnormal predictions in 124 of 375 cells and the ordinal model in 117 of 375 (mean 5.0 vs. 4.7 per patient, maximum 12; 2 vs. 4 patients judged entirely normal). These reductions directly address the domain-shift inflation of extreme predictions.

## 4. Discussion

### 4.1 Principal Findings

We developed a full-sequence attention-based MIL model with ordinal regression grading five conditions across five disc levels on sagittal T2-weighted MRI and validated it in a hospital cohort. Three findings stand out. First, full-sequence information is indispensable: the single-slice baseline achieved 16.3% balanced accuracy versus 51.4% for attention over 24 uniformly sampled sagittal slices (Table 2). Second, task difficulty is highly structured: Moderate was the hardest grade across every condition (AUC 0.70–0.78), consistent with documented inter-reader variability at the mild–moderate boundary [31], and performance across disc levels tracked prevalence (Spearman’s ρ = 0.8) — data scarcity at rarely affected levels (L1/L2, 4.2%), not model design, is the binding constraint. Third, the ordinal formulation changed decision behavior: severe-grade predictions contracted from 933 to 380 cells internally and from 172 to 69 externally (McNemar p<0.0001), without sacrificing sensitivity (23 vs. 29 false negatives). Attention analysis explains why: the highest-attention slices lay 5–8 slices from the sequence midpoint — precisely what a single-slice baseline discards.

### 4.2 Comparison with the Literature

Direct comparison with prior work is complicated by heterogeneous task definitions and metrics (Supplementary Table S1). The most comparable benchmark, the SpineNetV2-derived SGN, reports 79.4% balanced accuracy — for canal stenosis alone, dropping to 55.4% under direct transfer [9]. Multi-condition models remain rare: a preprint with five condition-specific vision transformers reported 82–92% validation accuracy and called for ordinal, patient-level evaluation [33]; a segmentation-based approach reported 94.9% accuracy under a non-official label scheme [34]. Our internal results (51.4% balanced accuracy; condition-averaged severe-grade AUC 0.908) reflect the true difficulty of the complete 25-task scheme, in line with competition solutions near 50–60% [13].

Externally, prior validations covered single conditions with non-standard labels, reported image-level metrics far above competition benchmarks (0.956 and 0.929), and provided no patient-level statistics [15,16]. To our knowledge, this is the first hospital external validation of the complete five-condition × five-level scheme with patient-level statistics (McNemar tests, patient-clustered bootstrap intervals) — the validation gap the segmentation study itself identified as the field’s outstanding need [34].

### 4.3 Why the Ordinal Formulation Matters

The ordinal-versus-weighted comparison isolates a behavioral rather than purely statistical effect: the variants differ only in the output layer (25×2 cumulative logits vs. 25×3 softmax heads) and achieve nearly identical ranking ability (Section 3.7), yet decide differently. Under domain shift, severe-grade false alarms reached 172 of 900 cells for the weighted model versus 69 for the ordinal (McNemar p<0.0001), without a cost in missed disease (23 vs. 29 false-negative cells). We attribute this to the cumulative-logit parameterization, which encodes grade ordering directly and penalizes implausible Normal/Mild↔Severe confusion — only 2.9% of true severe cells were misclassified as Normal/Mild. For a screening aid, false severe flags are disproportionately costly because they trigger unnecessary urgent referrals. The +3.5-point Moderate-grade AUC gain further suggests the ordinal bias helps precisely where the label boundary is most ambiguous.

### 4.4 Limitations

Our study has limitations. First, the external cohort is single-center and retrospective (60 patients, four vendors) with only seven severe ground-truth cells (0.8%); severe-grade recall is unreliable, and the balanced-accuracy reversal between models (0.395 vs. 0.468) should not be over-interpreted. Second, external ground truth came from clinical reports under a “not mentioned = normal” convention (25 of 60 reports mentioned no stenosis), which may bias prevalence estimates. Third, external validation covered only the three conditions routinely reported in Chinese practice (15 of 25 tasks). Fourth, the source dataset carries label noise from multiple radiologists, and the mild–moderate boundary is itself subjective [31]. Fifth, the final model discards axial information (the dual-plane variant underperformed sagittal-only input, 29.4% vs. 32.7%), so the axial plane’s potential remains unexplored. Sixth, absolute performance (51.4% balanced accuracy) limits the model to a screening/triage role rather than standalone diagnosis.

### 4.5 Clinical Implications and Future Directions

Two properties suggest a realistic deployment path: strong severe-grade discrimination across conditions (average AUC 0.908; 0.944 for canal stenosis) with rare gross errors (2.9% of severe cases called normal) — the properties of a rule-in triage screen — and interpretable attention concentrated on a small number of off-midline slices clinicians can inspect. Given the intrinsically ambiguous moderate grade, the tool should flag examinations for prioritized reading rather than issue standalone diagnoses. Future work should pursue multi-center prospective validation covering all five conditions, site-specific threshold calibration, and incorporation of the axial plane with slice-level registration; the ordering-based output may also facilitate structured reporting.

## 5. Conclusion

A full-sequence attention-based MIL model with ordinal regression achieved 51.4% balanced accuracy on the complete 25-task lumbar grading scheme of the LumbarDISC dataset, with a condition-averaged severe-grade AUC of 0.908. In the first hospital external validation of a model covering this complete scheme (60 patients, four scanner vendors), the ordinal model reduced severe-grade false alarms (69 vs. 172 predicted severe cells; McNemar p<0.0001) while preserving sensitivity — confirming that ordinal constraints mitigate the domain-shift inflation of extreme predictions.

## Data Availability

The public training dataset (RSNA 2024 LumbarDISC) is available online at https://www.kaggle.com/competitions/rsna-2024-lumbar-spine-degenerative-classification. All other data produced in the present study are available upon reasonable request to the authors.

## Acknowledgments

The authors thank the organizers of the RSNA 2024 Lumbar Spine Degenerative Classification challenge for the public LumbarDISC dataset. During the preparation of this manuscript, the authors used Claude (Anthropic) to assist with language editing; the authors reviewed and approved all output and take full responsibility for the content.

## Data Availability Statement

The RSNA 2024 Lumbar Spine Degenerative Classification (LumbarDISC) training dataset is publicly available (https://www.kaggle.com/competitions/rsna-2024-lumbar-spine-degenerative-classification). The external validation hospital imaging data are not publicly available owing to patient privacy; de-identified data may be made available by the corresponding author upon reasonable request and with institutional approval. The analysis code is available from the corresponding author upon reasonable request.

## Author contributions (CRediT)

Shijiang Geng: Conceptualization, Methodology, Software, Formal analysis, Investigation, Data curation, Writing – original draft, Visualization. Qizhuo Miao: Methodology, Validation, Data curation, Writing – review & editing. Qingqing Qin: Investigation, Resources, Data curation, Writing – review & editing. Haoran Guo: Investigation, Resources, Writing – review & editing. Xin Wang: Conceptualization, Methodology, Supervision, Project administration, Writing – review & editing.

---

**Supplementary Figure S1.**
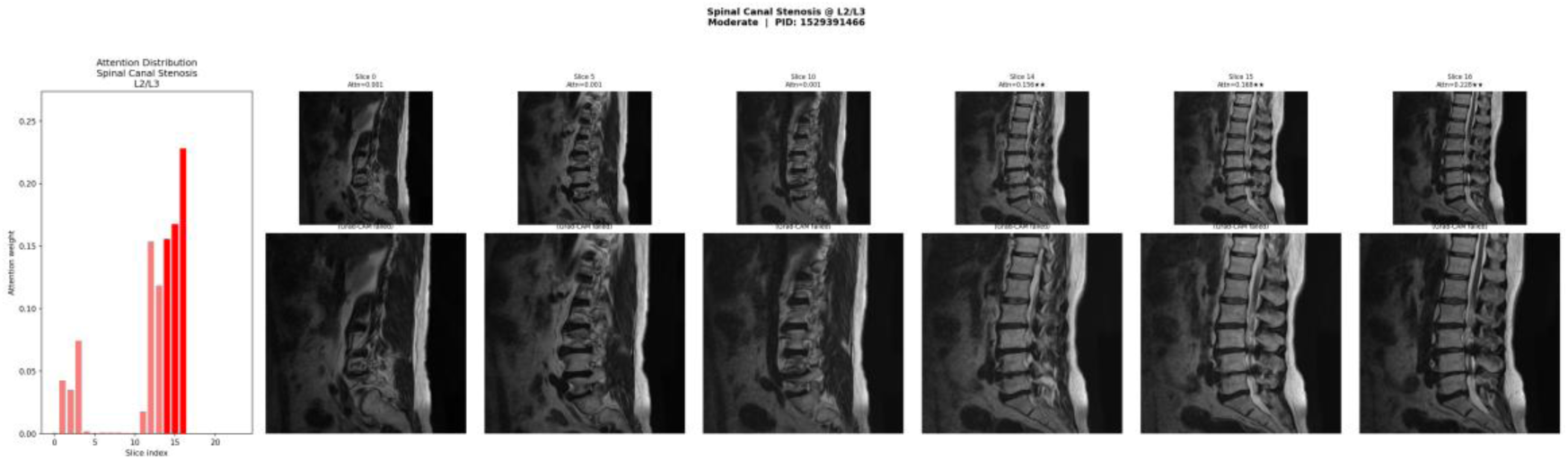
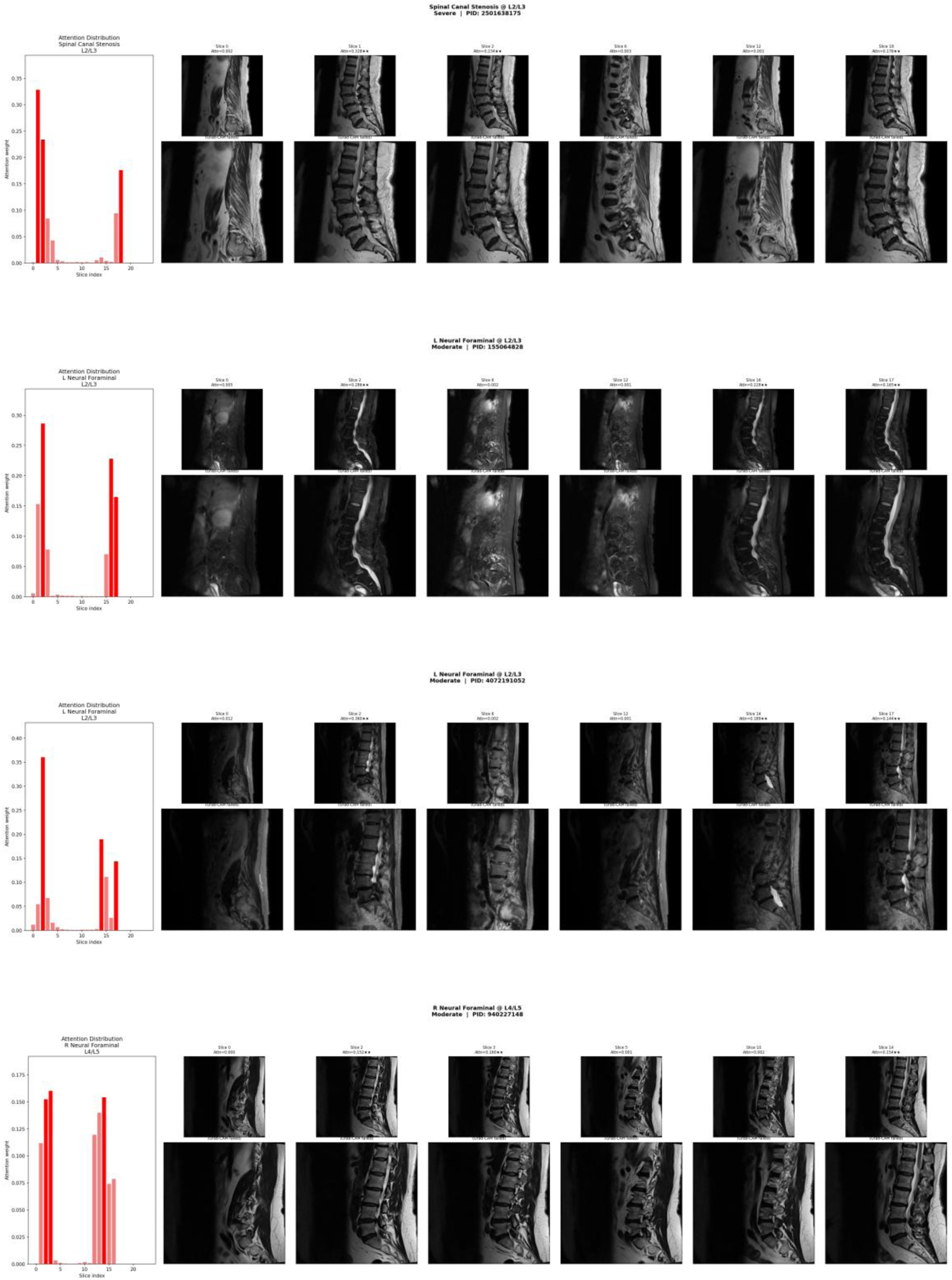
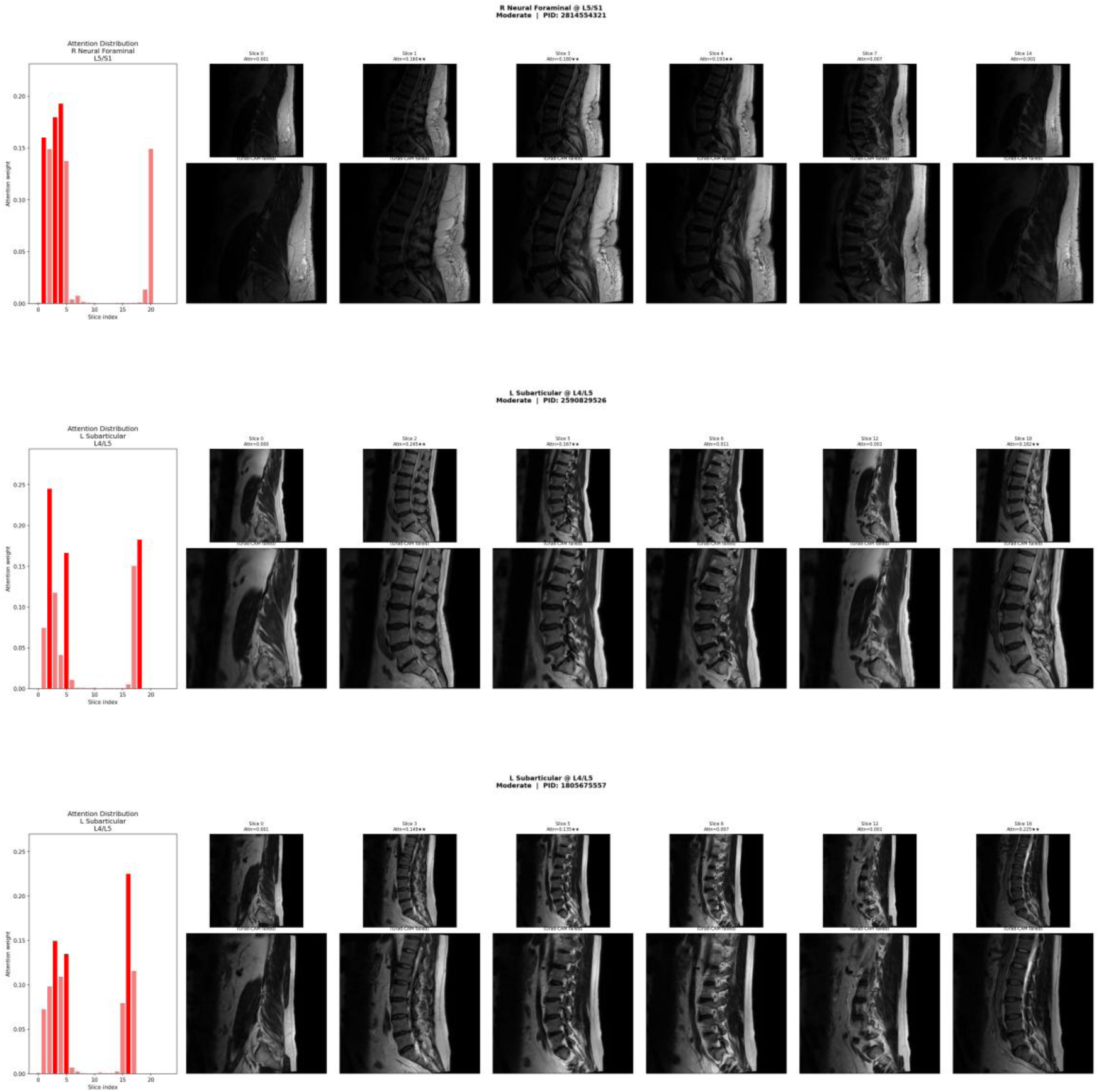
Grad-CAM heatmaps for eight representative cases, showing the image regions that drive the model’s predictions.

## Supplementary Materials

| Study (Year) | Architecture | Task Scope | Internal Result | External Validation | Notes |
| --- | --- | --- | --- | --- | --- |
| Ours (2026) | ResNet34 + biGRU + attention MIL, ordinal regression | 5 conditions × 5 levels × 3 grades (25 tasks) | Bal acc 51.4%; AUC 0.88/0.76/0.92 (NM/Mod/Sev) | 60 patients, multi-vendor, Chinese hospital; patient-level McNemar + clustered bootstrap CI | First hospital external validation of a complete-scheme model; 15 tasks reportable in Chinese practice |
| Jutzeler et al. (2026) | SGN (SpineNetV2 fine-tuned) | Canal stenosis only | Bal acc 79.4% (fine-tuned); 55.4% (direct transfer) | None | Benchmark study; transfer drop documents domain shift |
| Liu et al. (2025) | MobileNetV2-UNet segmentation + classification | Partial, non-official label scheme | Acc 94.9% (not directly comparable) | None | Authors call for hospital external validation |
| Trinc et al. (2026) | Five condition-specific ViTs (ViT-B/16, ViT-L/16, etc.) | 5 conditions × 5 levels × 3 grades | Val acc 82–92% | None | Preprint (not peer-reviewed); proposes ordinal, patient- level<br>evaluation<br>framework |
| Udomlu<br>ck et al.<br>(2026) | VGG19/ConvNeXt/DIN<br>Ov2 features +<br>LR/SVM/LightGBM | Canal<br>stenosis<br>only,<br>custom<br>4-level<br>labels | Acc 0.956 | 70<br>patients,<br>single<br>Thai<br>hospital,<br>no fine-tuning | Truly<br>independe<br>nt, but<br>single-<br>condition,<br>image-level, non-standard<br>labels |
| Hamid<br>et al.<br>(2026) | EfficientNet-B3 +<br>DenseNet-121 hybrid | Canal<br>stenosis<br>only | CV acc<br>97.4% | Banglades<br>hi hospital<br>(BSMMU<br>) | Conferenc<br>e paper<br>(IEEE);<br>not in<br>PubMed |
| **Disc Level** |  | **Proportion Abnormal** |  | **Balanced Accuracy** |  |
| L1/L2 |  | 4.2% |  | 26.3% |  |
| L2/L3 |  | 14.6% |  | 62.9% |  |
| L3/L4 |  | 27.4% |  | **75.6%** |  |
| **L4/L5** |  | **42.5%** |  | 72.6% |  |
| L5/S1 |  | 26.8% |  | 48.3% |  |

